# Analytical Validation of MyProstateScore 2.0

**DOI:** 10.1101/2024.08.06.24311421

**Authors:** Jacob I. Meyers, Tabea M. Schatz, Cameron J. Seitz, Rachel Botbyl, Bradley S. Moore, Bill G. Crafts, Spencer Heaton, John R. Kitchen

**Affiliations:** Lynx Dx, Ann Arbor, MI

**Keywords:** biomarkers, early detection of cancer, prostate cancer, liquid biopsy

## Abstract

Prostate cancer (PCa) remains the most common cancer and the second leading cause of cancer-related deaths among men in the United States. Early detection is critical, yet routine prostate-specific antigen (PSA) screening has shown limited benefits, often leading to unnecessary biopsies. MyProstateScore 2.0 (MPS2), a novel urinary 18-biomarker test, aims to improve the detection of clinically significant prostate cancer (csPCa) while minimizing the risk of unnecessary procedures. This study aims to validate the analytical performance of MPS2, including analytical detection limits, reproducibility, and robustness across various potential interfering substances. The limit of detection (LoD) and lower limit of quantitation (LLOQ) for the 18 MPS2 biomarkers ranged from 40 to 160 copies/reaction and 80 to 320 copies/reaction, respectively. The upper limit of quantitation (ULOQ) was determined to be up to 10^7 copies/reaction for most analytes. The linear amplification of MPS2 biomarker amplification was confirmed between the defined LLOQ and ULOQ. The impact of eleven potential interfering substances were tested and shown to have minimal impact on the assay. In conclusion, MPS2 exhibits robust analytical performance, with reliable detection and quantitation limits. This validation supports the clinical use of MPS2 for accurately predicting clinically-significant PCa, potentially reducing unnecessary biopsies and associated complications.

---

Prostate cancer (PCa) is the most common cancer and the second leading cause of cancer deaths for men in the United States.^1^ Survival is strongly associated with stage at diagnosis, with a 5-year relative survival rate of 96.1% for localized disease versus 32.3% for distant metastasized disease.^2^ This highlights the importance of early detection, however routine screening for PCa remains controversial. Prostate specific antigen (PSA) is the most commonly used biomarker for PCa screening, however routine PSA screening has a United States Preventive Services Task Force (USPSTF) rating of C for men 50-69 and D for men 70+, indicating unclear benefit.^3^ PSA screening has shown little to no improvement in PCa mortality^4^ and is associated with increased harm resulting from unnecessary prostate biopsies.^5-6^ Thus, there is a need for a PCa screening test that improves early detection of clinically significant PCa while minimizing the risk of unnecessary biopsy and associated complications.

Urinary biomarkers have emerged as a promising method for improved detection of clinically significant prostate cancer (csPCa).^7^ Among these is MyProstateScore 2.0 (MPS2), a novel PCa risk prediction test which utilizes 18 prostate-cancer-specific biomarkers to improve predictive accuracy for PCa with Grade Group 2 or higher (GG≥2). The test performed well in a clinical validation study with negative predictive values ranging from 95% to 99% for csPCa thus providing superior diagnostic accuracy to previously available urinary biomarker tests.^8^

This study aims to validate the analytical performance of MPS2, including determination of MPS2 analyte detection limits, reproducibility across relevant technical parameters, and robustness in the presence of potential interfering substances.

## MATERIALS AND METHODS

### RNA Extraction, Reverse Transcription, Pre-Amplification and Quantitative PCR

RNA was extracted from 0.5 mL of digital rectal exam (DRE) urine (ThermoFisher Scientific, Cat. No. A27828) using a customized protocol on a KingFisher™ Flex Purification System. Extracted RNA was reverse transcribed into complementary DNA (cDNA) (ThermoFisher Scientific, Cat. No. 11756050). Following reverse transcription, MPS2 biomarkers were pre-amplified for 14 cycles with biomarker-specific oligos and random hexamers (ThermoFisher Scientific, Cat. No. 4391128). Pre-amplified DNA was diluted and used as a template for Quantitative Polymerase Chain Reaction (qPCR). qPCR was performed on the OpenArray™ System using a QuantStudio™ 12K Flex Accufill System and custom OpenArray plates containing primers and probes for the 18 MPS2 biomarkers.

### Determination of Limit of Detection and Lower Limit of Quantitation

Eight concentrations (320, 160, 80, 40, 20, 10, 2.5, 1.25 copies/reaction) of a pool containing 18 artificially synthesized MPS2 biomarkers were used to determine the limit of detection (LoD) and the lower limit of quantitation (LLOQ) for each analyte. For both LOD and LLOQ, at least 16 replicates were run of each concentration across 12 runs. The LOD for each analyte was defined as the lowest concentration that could be detected in ≥95% of the replicates. The LLOQ for each analyte was defined as the lowest concentration with a standard deviation ≤0.7 cycle threshold (Crt), which is under the previously defined acceptance criteria of standard deviation under 1.0 Crt.^9-10^

### Determination of Linear Range and Upper Limit of Quantitation

Six concentrations (10^7^, 10^6^, 10^5^, 10^4^, 10^3^, and 10^2^ copies/reaction) of a pool containing 18 artificially synthesized MPS2 biomarkers were used to determine the linear range and the upper limit of quantitation (ULOQ) of each MPS2 analyte. Eight replicates of each concentration were run for a total of 48 results. The ULOQ was defined as the highest concentration with a standard deviation under 0.5 Crt. The linear range was defined as the range of concentrations within the ULOQ and LLOQ with a qPCR efficiency of 95% to 105% and R^2^>0.975.

### Precision

MPS2 precision was measured across several variables including equipment (QuantStudio™ 12K Flex Accufill Systems, and VeritiPro), operator (Tech A and Tech B), intra-run (repeatability), and inter-run (reproducibility). All precision experiments were performed using a pool of all 18 artificially synthesized MPS2 analytes. Three concentrations of that pool (3200, 1600, 800 copies/reaction) with eight replicates of each were tested on two VeritiPro thermocyclers and two QuantStudio™ 12K Flex Accufill Systems by two technicians over three days.

The criterion for passing precision was a standard deviation under 0.5 Crt for each MPS2 analyte across replicates.

### Interfering Substances

To determine whether certain substances interfere with the assay, 11 potential interfering substances were added to urine. The 11 substances and the concentrations tested are shown in Table 4. Contrived samples were created by adding artificially synthesized MPS2 analytes to artificial urine (Flinn Scientific, Catalog Number FB1444) to mimic a clinical urine sample. Note, this artificial urine is synthetically manufactured to mimic human urine and does not include human-derived substances. This allowed complete control over the concentration of substances normally found in human urine. For each substance, six replicates were tested as well as at least four replicates of the contrived urine sample without any substance added. The established criterion for passing is a change in the normalized Crt ≤ 0.5.

## RESULTS

### Limit of Detection and Lower Limit of Quantitation

The LoD and LLOQ were determined by testing eight concentrations ranging from 320 to 1.25 copies/reaction. The LoD for each analyte was defined as the lowest concentration that could be detected in ≥95% of the replicates. The LoD for the 18 analytes ranges from 40 to 160 copies per reaction (Table 1). The LoD of CAMKK2, KLK4, PCGEM1, SPON2, TFF3, TMSB15A and TRGV9 was determined to be 40 copies/reaction with 56 replicates. The LoD of APOC1, B3GNT6, ERG, HOXC6, NKAIN1, OR51E2, PCA3, PCAT14 and T2ERG was determined to be 80 copies/reaction while the LoD of KLK3 and SCHLAP1 was determined to be 160 copies/reaction.

**Table 1.** MPS2 analyte limit of detection (LoD), lower limit of quantitation (LLOQ) and upper limit of quantitation (ULOQ)

| MPS2 analyte | LOD in copies/reaction<br>(% detection) | LLOQ in copies/reaction (SD) | ULOQ in copies/reaction (SD) |
| --- | --- | --- | --- |
| APOC1 | 80 (100%) | 160 (0.43) | 10,000,000 (0.17) |
| B3GNT6 | 80 (100%) | 160 (0.53) | 10,000,000 (0.08) |
| CAMKK2 | 40 (95%) | 160 (0.45) | 10,000,000 (0.08) |
| ERG | 80 (97%) | 80 (0.69) | 10,000,000 (0.06) |
| HOXC6 | 80 (100%) | 320 (0.43) | 10,000,000 (0.07) |
| KLK3 | 160 (100%) | 160 (0.53) | 10,000,000 (0.06) |
| KLK4 | 40 (100%) | 320 (0.40) | 1,000,000 (0.34) |
| NKAIN1 | 80 (97%) | 80 (0.59) | 10,000,000 (0.16) |
| OR51E2 | 80 (100%) | 320 (0.27) | 10,000,000 (0.13) |
| PCA3 | 80 (100%) | 320 (0.33) | 10,000,000 (0.10) |
| PCAT14 | 80 (98%) | 160 (0.45) | 10,000,000 (0.12) |
| PCGEM1 | 40 (95%) | 160 (0.46) | 10,000,000 (0.11) |
| SCHLAP1 | 160 (100%) | 160 (0.74) | 10,000,000 (0.12) |
| SPON2 | 40 (95%) | 80 (0.60) | 10,000,000 (0.06) |
| TFF3 | 40 (95%) | 160 (0.37) | 10,000,000 (0.05) |
| T2ERG | 80 (100%) | 160 (0.69) | 10,000,000 (0.04) |
| TMSB15A | 40 (95%) | 160 (0.53) | 10,000,000 (0.08) |
| TRGV9 | 40 (96%) | 160 (0.35) | 10,000,000 (0.12) |

The LLOQ for each analyte was defined as the lowest concentration with a standard deviation ≤0.7 Crt. The LLOQ of ERG, NKAIN1, and SPON2 was determined to be 80 copies/reaction with 32 replicates. The LLOQ of APOC1, B3GNT6, CAMKK2, KLK3, PCAT14, PCGEM1, SCHLAP1, TFF3, T2ERG, TMSB15A, and TRGV9 was determined to be 160 copies/reaction with 16 replicates. The LLOQ of HOXC6, KLK4, OR51E2, and PCA3 was determined to be 320 copies/reaction with 16 replicates.

### Upper Limit of Quantitation and Linear Range

The ULOQ was defined as the highest concentration with a standard deviation under 0.5 Crt (Table 1). Most of the analytes (17/18) were found to have a ULOQ of 10^7^ copies/reaction. The ULOQ of KLK4 was determined to be 10^6^ copies per reaction. The lower ULOQ for KLK4 was due to template oversaturation, causing no amplification to occur.

The linear range and ULOQ were determined by testing six concentrations (10^7^, 10^6^, 10^5^, 10^4^, 10^3^, and 10^2^ copies/reaction). The linear range was defined as the range of concentrations within the ULOQ and LLOQ with a qPCR efficiency of 95% to 105% and R^2^>0.975. All MPS2 analytes exhibited linear amplification between their individually determined LLOQ and ULOQ with qPCR efficiencies ranging from 97% to 105% and R^2^ values ranging from 0.98 to 1.00 (Figure 1, Table 2).

**Table 2.** Linear amplification of MPS2 analytes.

| Target | R<sup>2</sup> | PCR Efficiency (%) |
| --- | --- | --- |
| APOC1 | 0.9997 | 98% |
| B3GNT6 | 0.9986 | 101% |
| CAMKK2 | 0.999 | 102% |
| ERG | 0.999 | 101% |
| HOXC6 | 0.9994 | 102% |
| KLK3 | 0.9991 | 100% |
| KLK4 | 1 | 99% |
| NKAIN1 | 0.9985 | 105% |
| OR51E2 | 0.9992 | 101% |
| PCA3 | 0.9991 | 101% |
| PCAT14 | 0.9995 | 101% |
| PCGEM1 | 0.98 | 98% |
| SCHLAP1 | 0.9994 | 98% |
| SPON2 | 0.9998 | 97% |
| TFF3 | 0.9988 | 102% |
| T2ERG | 0.998 | 98% |
| TMSB15A | 0.9994 | 100% |
| TRGV9 | 0.9997 | 100% |

**Figure 1.**
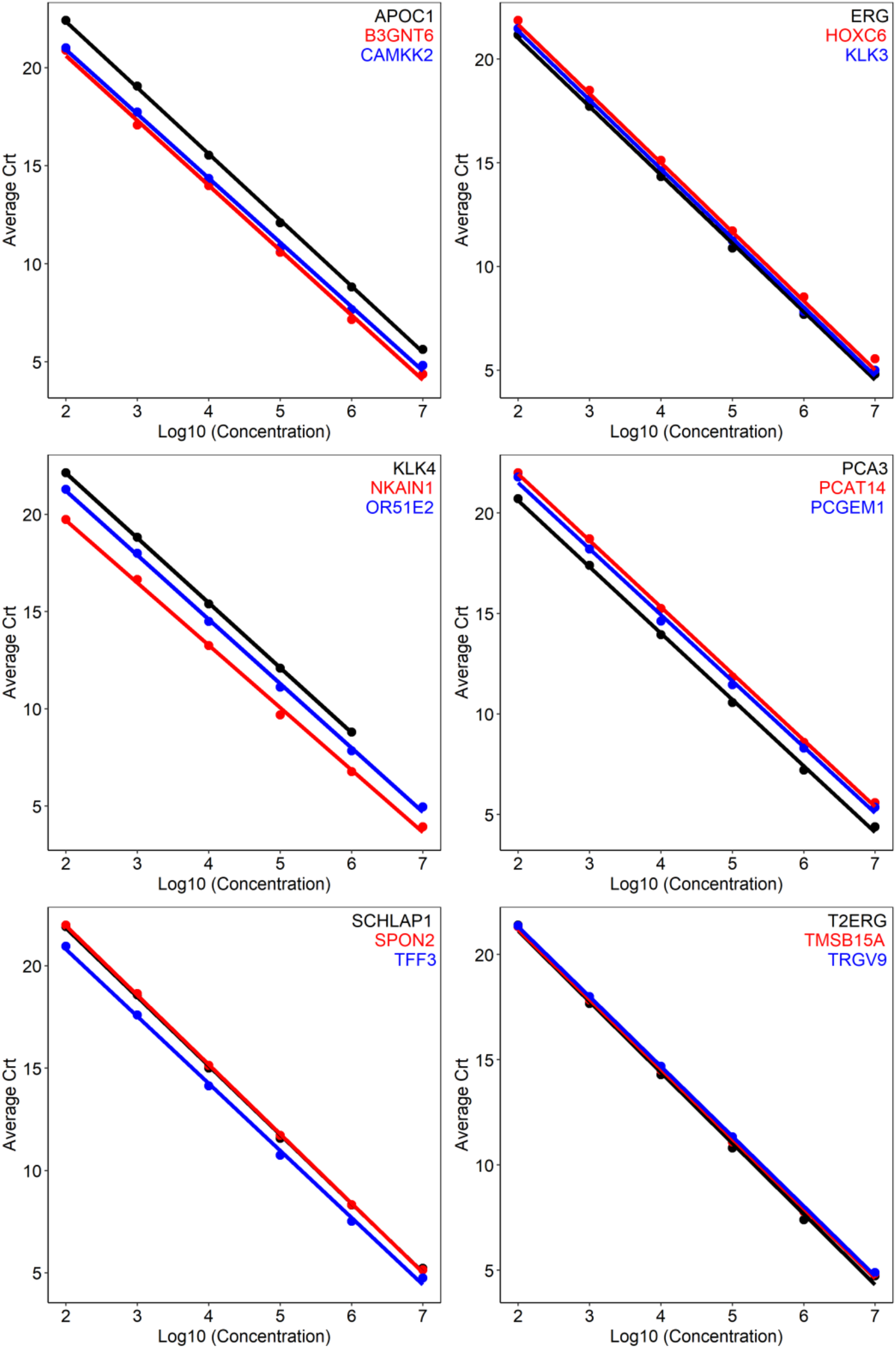
Linear regression of MPS2 analyte amplification.

### Precision

Three concentrations (3200, 1600, 800 copies/reaction) of a pool of the 18 artificially synthesized MPS2 biomarkers were run by two technicians over three days using two sets of equipment. Eight replicates of each concentration yielded a total of 143 valid results. From this dataset overall precision, repeatability, reproducibility, inter-instrument, and inter-technician precision were analyzed. The acceptance criterion for all precision was a standard deviation under 0.5 Crt, which passed for each MPS2 analyte (Table 3).

**Table 3.** Precision of MPS2 analyte detection.

| Target | Copies/reaction | Overall precision | Intra-run<br>(repeatability) | Inter-run<br>(reproducibility) | Inter-instrument<br>(QuantStudio™ 12K<br>Flex Accufill<br>System) | Inter-instrument<br>(VeritiPro) | Inter-technician |
| --- | --- | --- | --- | --- | --- | --- | --- |
| APOC1 | 3200 | 16.6+/-0.2 | 16.6+/-0.2 | 16.7+/-0.2 | 16.5+/-0.2 | 16.7+/-0.1 | 16.7+/-0.2 |
|  | 1600 | 17.8+/-0.2 | 17.9+/-0.1 | 17.9+/-0.2 | 17.9+/-0.1 | 17.8+/-0.2 | 17.8+/-0.2 |
|  | 800 | 18.8+/-0.3 | 18.8+/-0.2 | 18.9+/-0.2 | 18.8+/-0.2 | 18.8+/-0.3 | 18.9+/-0.2 |
| B3GNT6 | 3200 | 15.5+/-0.2 | 15.3+/-0.1 | 15.6+/-0.2 | 15.3+/-0.1 | 15.6+/-0.2 | 15.5+/-0.3 |
|  | 1600 | 16.7+/-0.2 | 16.7+/-0.1 | 16.8+/-0.1 | 16.7+/-0.1 | 16.7+/-0.2 | 16.8+/-0.1 |
|  | 800 | 17.6+/-0.3 | 17.6+/-0.2 | 17.8+/-0.2 | 17.5+/-0.2 | 17.6+/-0.3 | 17.7+/-0.3 |
| CAMKK2 | 3200 | 15.7+/-0.2 | 15.5+/-0.1 | 15.7+/-0.2 | 15.5+/-0.1 | 15.7+/-0.2 | 15.7+/-0.2 |
|  | 1600 | 16.8+/-0.2 | 16.8+/-0.1 | 16.9+/-0.1 | 16.8+/-0.1 | 16.9+/-0.2 | 16.9+/-0.1 |
|  | 800 | 17.8+/-0.2 | 17.7+/-0.2 | 17.9+/-0.2 | 17.7+/-0.1 | 17.8+/-0.2 | 17.9+/-0.2 |
| ERG | 3200 | 15.7+/-0.2 | 15.6+/-0.1 | 15.8+/-0.2 | 15.6+/-0.1 | 15.8+/-0.2 | 15.7+/-0.2 |
|  | 1600 | 16.9+/-0.2 | 16.9+/-0.1 | 17+/-0.1 | 16.9+/-0.1 | 17+/-0.2 | 17+/-0.1 |
|  | 800 | 17.8+/-0.3 | 17.8+/-0.2 | 18+/-0.2 | 17.8+/-0.1 | 17.9+/-0.3 | 18+/-0.2 |
| HOXC6 | 3200 | 16.3+/-0.2 | 16.2+/-0.2 | 16.4+/-0.2 | 16.1+/-0.2 | 16.4+/-0.2 | 16.3+/-0.2 |
|  | 1600 | 17.6+/-0.2 | 17.6+/-0.1 | 17.6+/-0.1 | 17.6+/-0.1 | 17.6+/-0.2 | 17.6+/-0.1 |
|  | 800 | 18.4+/-0.3 | 18.5+/-0.2 | 18.6+/-0.2 | 18.4+/-0.2 | 18.5+/-0.3 | 18.6+/-0.2 |
| KLK3 | 3200 | 16+/-0.2 | 15.8+/-0.1 | 16.1+/-0.2 | 15.8+/-0.1 | 16.1+/-0.2 | 16+/-0.2 |
|  | 1600 | 17.3+/-0.2 | 17.3+/-0.2 | 17.4+/-0.1 | 17.3+/-0.2 | 17.3+/-0.2 | 17.4+/-0.2 |
|  | 800 | 18.1+/-0.3 | 18.2+/-0.2 | 18.3+/-0.2 | 18.1+/-0.2 | 18.2+/-0.3 | 18.3+/-0.2 |
| KLKL4 | 3200 | 16.7+/-0.2 | 16.6+/-0.2 | 16.7+/-0.1 | 16.5+/-0.2 | 16.7+/-0.1 | 16.7+/-0.2 |
|  | 1600 | 18+/-0.2 | 18+/-0.1 | 18+/-0.1 | 18+/-0.1 | 18+/-0.2 | 18+/-0.1 |
|  | 800 | 18.9+/-0.3 | 18.9+/-0.3 | 19.1+/-0.2 | 18.9+/-0.3 | 19+/-0.3 | 19+/-0.3 |
| NKAIN1 | 3200 | 14.5+/-0.2 | 14.4+/-0.3 | 14.4+/-0.2 | 14.3+/-0.2 | 14.5+/-0.2 | 14.4+/-0.2 |
|  | 1600 | 15.7+/-0.2 | 15.8+/-0.1 | 15.7+/-0.1 | 15.8+/-0.1 | 15.7+/-0.2 | 15.7+/-0.1 |
|  | 800 | 16.5+/-0.3 | 16.6+/-0.2 | 16.7+/-0.2 | 16.6+/-0.2 | 16.5+/-0.3 | 16.6+/-0.2 |
| OR51E2 | 3200 | 15.7+/-0.1 | 15.6+/-0.1 | 15.7+/-0.1 | 15.6+/-0.1 | 15.7+/-0.1 | 15.7+/-0.1 |
|  | 1600 | 16.9+/-0.1 | 17+/-0.1 | 17+/-0.1 | 17+/-0.1 | 17+/-0.1 | 17+/-0.1 |
|  | 800 | 17.8+/-0.2 | 17.9+/-0.2 | 18+/-0.2 | 17.8+/-0.2 | 17.9+/-0.2 | 17.9+/-0.2 |

**Table 3 (continued).** Precision of MPS2 analyte detection
| Target | Copies/reaction | Overall precision | Intra-run<br>(repeatability) | Inter-run<br>(reproducibility) | Inter-instrument<br>(QuantStudio™ 12K<br>Flex Accufill<br>System) | Inter-instrument<br>(VeritiPro) | Inter-technician |
| --- | --- | --- | --- | --- | --- | --- | --- |
| PCA3 | 3200 | 15.5+/-0.2 | 15.3+/-0.2 | 15.6+/-0.2 | 15.3+/-0.2 | 15.5+/-0.2 | 15.5+/-0.2 |
|  | 1600 | 16.7+/-0.2 | 16.7+/-0.1 | 16.8+/-0.1 | 16.7+/-0.1 | 16.7+/-0.2 | 16.8+/-0.1 |
|  | 800 | 17.7+/-0.3 | 17.7+/-0.2 | 17.9+/-0.2 | 17.7+/-0.2 | 17.7+/-0.3 | 17.8+/-0.2 |
| PCAT14 | 3200 | 16.6+/-0.2 | 16.4+/-0.1 | 16.6+/-0.2 | 16.5+/-0.1 | 16.6+/-0.2 | 16.6+/-0.2 |
|  | 1600 | 17.8+/-0.2 | 17.8+/-0.1 | 17.8+/-0.1 | 17.8+/-0.1 | 17.8+/-0.2 | 17.8+/-0.1 |
|  | 800 | 18.7+/-0.2 | 18.6+/-0.2 | 18.8+/-0.2 | 18.6+/-0.2 | 18.7+/-0.2 | 18.8+/-0.2 |
| PCGEM1 | 3200 | 16.4+/-0.2 | 16.2+/-0.1 | 16.5+/-0.2 | 16.2+/-0.1 | 16.4+/-0.2 | 16.4+/-0.2 |
|  | 1600 | 17.6+/-0.2 | 17.7+/-0.1 | 17.7+/-0.1 | 17.7+/-0.1 | 17.6+/-0.2 | 17.7+/-0.1 |
|  | 800 | 18.5+/-0.3 | 18.5+/-0.2 | 18.7+/-0.2 | 18.5+/-0.2 | 18.6+/-0.3 | 18.7+/-0.3 |
| SCHLAP1 | 3200 | 16.3+/-0.2 | 16.2+/-0.1 | 16.4+/-0.2 | 16.2+/-0.1 | 16.4+/-0.2 | 16.3+/-0.2 |
|  | 1600 | 17.6+/-0.2 | 17.6+/-0.1 | 17.7+/-0.1 | 17.6+/-0.1 | 17.6+/-0.2 | 17.6+/-0.1 |
|  | 800 | 18.4+/-0.3 | 18.4+/-0.2 | 18.6+/-0.2 | 18.4+/-0.2 | 18.5+/-0.3 | 18.5+/-0.2 |
| SPON2 | 3200 | 16.6+/-0.2 | 16.4+/-0.1 | 16.6+/-0.2 | 16.4+/-0.2 | 16.6+/-0.2 | 16.6+/-0.2 |
|  | 1600 | 17.8+/-0.2 | 17.9+/-0.1 | 17.9+/-0.2 | 17.8+/-0.1 | 17.9+/-0.2 | 17.9+/-0.2 |
|  | 800 | 18.8+/-0.3 | 18.8+/-0.2 | 19+/-0.2 | 18.8+/-0.2 | 18.9+/-0.3 | 19+/-0.3 |
| TFF3 | 3200 | 15.6+/-0.1 | 15.5+/-0.1 | 15.6+/-0.2 | 15.5+/-0.1 | 15.6+/-0.2 | 15.6+/-0.2 |
|  | 1600 | 16.8+/-0.2 | 16.8+/-0.1 | 16.8+/-0.1 | 16.8+/-0.1 | 16.8+/-0.2 | 16.9+/-0.1 |
|  | 800 | 17.7+/-0.2 | 17.7+/-0.2 | 17.8+/-0.2 | 17.6+/-0.2 | 17.7+/-0.2 | 17.8+/-0.2 |
| T2ERG | 3200 | 15.4+/-0.2 | 15.1+/-0.2 | 15.4+/-0.3 | 15.2+/-0.2 | 15.4+/-0.2 | 15.4+/-0.3 |
|  | 1600 | 16.6+/-0.2 | 16.6+/-0.2 | 16.7+/-0.1 | 16.6+/-0.1 | 16.6+/-0.2 | 16.7+/-0.2 |
|  | 800 | 17.5+/-0.3 | 17.4+/-0.2 | 17.7+/-0.2 | 17.4+/-0.2 | 17.6+/-0.3 | 17.6+/-0.3 |
| TMSB15A | 3200 | 15.8+/-0.2 | 15.7+/-0.1 | 15.9+/-0.2 | 15.7+/-0.1 | 15.9+/-0.2 | 15.8+/-0.2 |
|  | 1600 | 17.1+/-0.2 | 17.1+/-0.1 | 17.2+/-0.1 | 17.1+/-0.1 | 17.1+/-0.2 | 17.2+/-0.2 |
|  | 800 | 17.9+/-0.3 | 17.9+/-0.2 | 18.1+/-0.2 | 17.9+/-0.2 | 18+/-0.3 | 18+/-0.3 |
| TRGV9 | 3200 | 16+/-0.2 | 15.9+/-0.1 | 16.1+/-0.2 | 15.9+/-0.1 | 16.1+/-0.2 | 16+/-0.2 |
|  | 1600 | 17.3+/-0.2 | 17.3+/-0.2 | 17.4+/-0.2 | 17.3+/-0.2 | 17.3+/-0.2 | 17.4+/-0.2 |
|  | 800 | 18.2+/-0.3 | 18.1+/-0.3 | 18.4+/-0.3 | 18.1+/-0.2 | 18.2+/-0.3 | 18.3+/-0.3 |

**Table 4.** MPS2 interfering substances testing.

| Substance | Conc. | APOC<br>1 | B3GN<br>T6 | CAMK<br>K2 | ERG | HOXC<br>6 | KLK3 | KLK4 | NKAIN<br>1 | OR51<br>E2 | PCA3 | PCAT1<br>4 | PCGE<br>M1 | SCHL<br>AP1 | SPON<br>2 | TFF3 | T2ER<br>G | TMSB<br>15A | TRGV<br>9 |
| --- | --- | --- | --- | --- | --- | --- | --- | --- | --- | --- | --- | --- | --- | --- | --- | --- | --- | --- | --- |
| Alcohol (100% ethanol) | 2.50% | -0.1 | -0.5 | -0.1 | -0.1 | 0.1 | 0 | 0 | -0.3 | -0.3 | -0.2 | -0.2 | -0.2 | -0.1 | -0.2 | -0.3 | 0.1 | 0 | 0 |
| Triglycerides | 15,000 mg/L | -0.1 | -0.3 | -0.1 | -0.1 | -0.1 | 0 | -0.4 | 0 | -0.1 | 0 | 0.1 | 0 | -0.1 | 0 | -0.1 | 0 | 0.1 | 0.1 |
| Hemoglobin | 2,000 mg/L | -0.1 | -0.1 | -0.1 | -0.1 | 0 | 0 | -0.1 | 0.1 | 0 | 0 | 0.2 | -0.1 | 0 | -0.1 | -0.2 | 0.3 | 0.3 | 0.1 |
| Calcium (CaCl) | 800 mg/L | -0.1 | -0.3 | -0.2 | 0 | -0.1 | 0 | -0.2 | 0.2 | 0 | -0.1 | 0.1 | 0 | 0.2 | -0.1 | -0.1 | 0.1 | 0.3 | 0.1 |
| Cholesterol | 700 mg/L | -0.4 | -0.4 | -0.2 | -0.1 | 0 | 0 | -0.3 | -0.1 | -0.4 | 0 | 0 | -0.2 | -0.2 | -0.1 | -0.1 | 0.1 | 0 | 0 |
| Glucose | 400 mg/L | -0.2 | -0.3 | -0.1 | -0.1 | 0 | 0 | -0.1 | -0.1 | -0.2 | 0 | 0 | -0.1 | -0.3 | 0.2 | 0 | 0.1 | -0.1 | 0.1 |
| Sodium (NaCl) | 8,000 mg/L | -0.2 | -0.2 | 0 | 0.2 | 0.2 | 0 | 0 | 0 | 0 | 0.2 | 0.3 | 0 | 0.2 | 0.1 | 0 | -0.3 | 0.2 | 0.3 |
| Albumin | 400 mg/L | -0.3 | -0.1 | 0.2 | -0.1 | -0.1 | 0 | -0.1 | 0.1 | 0.3 | 0.3 | 0.3 | 0.1 | -0.3 | 0.1 | 0.3 | 0.4 | 0.2 | 0.2 |
| Microorganisms | 257 cfu/mL | -0.1 | -0.1 | 0 | 0.1 | 0 | 0 | -0.1 | 0.1 | 0.1 | 0 | 0.1 | 0 | -0.1 | 0 | -0.1 | -0.2 | 0 | 0.3 |
| Free Bilirubin | 60 mg/L | -0.1 | -0.3 | -0.1 | -0.1 | 0.1 | 0 | -0.2 | -0.2 | -0.1 | -0.1 | 0 | -0.1 | -0.1 | 0 | -0.1 | 0.2 | 0.2 | 0.4 |
| Total Proteins | 1 mg/L | -0.3 | -0.2 | -0.2 | -0.1 | -0.1 | 0 | -0.1 | -0.1 | -0.3 | -0.1 | 0 | 0 | -0.1 | 0 | -0.1 | 0.1 | 0.1 | -0.1 |

Abbreviations: Conc., Concentration.
Measurements presented as change in normalized Crt (control – substance).

Intra-run (repeatability) was determined by one experiment run by one technician using one set of equipment over 24 valid results. Intra-run precision passed with a standard deviation ranging from 0.1 to 0.3 Crt which was less than the cut off 0.5 Crt (Table 3).

Inter-run (reproducibility) precision was determined by three experiments run by one technician over three days using one set of equipment over 71 valid results. Inter-run precision passed with a standard deviation ranging from 0.1 to 0.3 Crt which was less than the cut off 0.5 Crt (Table 3).

Inter-instrument (QuantStudio™ 12K Flex Accufill System) precision was determined by two experiments run by one technician over two days using two different QuantStudio™ 12K Flex Accufill Systems across 48 valid results. Inter-instrument (QuantStudio™ 12K Flex Accufill System) precision passed with a standard deviation ranging from 0.1 to 0.3 Crt which was less than the cut off 0.5 Crt (Table 3).

Inter-instrument (VeritiPro) precision was determined by three experiments run using two different VeritiPro thermocyclers across 95 valid results. Inter-instrument (VeritiPro) precision passed with a standard deviation ranging from 0.1 to 0.3 Crt (Table 4).

Inter-technician precision was determined by two experiments run by two technicians over two days using the same equipment across 48 valid results. Inter-technician precision passed with a standard deviation ranging from 0.1 to 0.3 Crt (Table 3).

### Interfering Substances

Eleven substances were tested to determine if they interfered with MPS2 analyte detection. This included 9 endogenous substances and 2 exogenous substances (alcohol, microorganisms). All substances were determine to have no interference with MPS2, with ≤0.5 normalized Crt deviation compared to controls.

## DISCUSSION

In this study, we comprehensively evaluated the ability of MPS2 to reproducibly measure target MPS2 analytes. The limits of MPS2 analytes were analytically determined based on Clinical & Laboratory Standards Institute (CLSI) guidelines and analytical validations performed on similarly designed prostate cancer diagnostic screening tests.^9, 11-14^ The LoD for MPS2 analytes were in a similar range as other urine-based prostate cancer screening tests, with the lower LoD of MPS2 analytes ranging from 40 to 160 copies per reaction and an ULOQ between 10^5^ and 10^7^ copies per reaction.^15-16^ MPS2 analyte detection in this range is highly linear (R^2^≥0.98) with PCR efficiencies ranging from 98% to 105%, indicating minimal to no amplification bias.

The robustness and reproducibility of MPS2 analyte detection was measured across time, technicians, pre-amplification thermal cyclers (VeritiPro) and real-time PCR systems (QuantStudio12K Flex OpenArray System). MPS2 analyte signals were highly reproducible across these factors at three concentrations, illustrating the robustness of MPS2 for clinical testing.

Finally, the impact of eleven substances that could occur in clinical urine samples was tested on MPS2 analyte detection. All substances were tested at elevated concentrations compared to what would typically be observed in human urine. The impact of eleven substances were tested on MPS2 analyte detection. None of the substances at the concentrations tested affected MPS2 performance.

## CONCLUSION

The study confirms the robustness of MPS2 to accuracy and reproducibly detect the target analytes. In conjunction with the previous clinical validation,^8^ this work further validates the biomarker signals measured in MPS2 are accurate and can be used clinically to predict clinically significant prostate cancer.

## Data Availability

All data produced in the present study are available from the authors upon reasonable request and with permission of Lynx Dx.

## Abbreviations and Acronyms

Crt: Cycle Threshold
csPCa: Clinically Significant Prostate Cancer
LLOQ: Lower Limit of Quantitation
LoD: Limit of Detection
MPS2: MyProstateScore 2.0
PCa: Prostate Cancer
PSA: Prostate-Specific Antigen
qPCR: Quantitative Polymerase Chain Reaction
RNase: Ribonuclease
ULOQ: Upper Limit of Quantitation

